# DNA Methylation Study in Presbycusis Patients

**DOI:** 10.1101/2022.10.31.22281760

**Authors:** Marie Valerie Roche, Denise Yan, Dana Godrich, Naser Hamad, Pei-Ciao Tang, Juan Young, Susan Blanton, Feng Gong, Xue Zhong Liu

**Affiliations:** Department of Otolaryngology, University of Miami Miller School of Medicine, Miami FL 33136, USA; Department of Biochemistry and Molecular Biology, University of Miami Miller School of Medicine, Miami, FL 33136, US; John P. Hussman Institute for Human Genomics; John T Macdonald Foundation Department of Human Genetics, University of Miami Miller School of Medicine, Miami, FL 33136

**Author notes:** <u>Corresponding Author:</u> Xue Zhong Liu, M.D., Ph.D. Professor of Otolaryngology, University of Miami Miller School of Medicine, Address: 1120 NW 14^th^ Street, 5^th^ Floor, Miami, FL 33136.

**Keywords:** Epigenetics, Presbycusis, Inner Ear, Cpg sites methylation, Hearing Loss, DNA Methylation, Methylation-Specific PCR

## Abstract

**Background:** Presbycusis, also known as age-related hearing loss (ARHL), is the most frequent sensory disability affecting elderly adults worldwide.ARHL is typified by a bilateral, progressive, sensorineural hearing loss that is pronounced in high frequency. Conventional factors associated with ARHL include diabetes, hypertension, and family history of hereditary hearing loss. The severity of hearing impairment varies between individuals. The accurate causative molecular pathogenesis for ARHL is unknown, therefore the investigation of the underlying pathogenic mechanisms involved in ARHL is imperative for the development of effective therapeutic approaches. Epigenetics is the study of phenotypic changes caused by modification of genetic expression rather than alteration of DNA sequence. It is hypothesized that ARHL could result from unclarified epigenetic susceptibility, nevertheless, there is a shortage of information on the exact contribution of epigenetic modifications to ARHL. Here we present an investigation on the involvement of DNA methylation with Age-related hearing loss.

**Results:** In the present study the Illumina Infinium® Methylation EPIC Beadchip has been used to identify regions with aberrant levels of methylation across genomes from ARHL patients. Hearing measurements were used to determine the audioprofiles. Clinical, audiometric patterns, DNA testing, and methylation pattern screening were undertaken. Our results demonstrate a strong correlation between patients’ hearing measurements and CpG sites methylation in *ESPN* and *TNFRSF25*. A Methylation Polymerase chain reaction (PCR) assay was used to confirm methylation levels at specific gene locus in ARHL patients.

**Conclusion:** Aberrant DNA methylation and its impact on gene expression have been implicated in many biological processes. By interrogating methylation status across the genome at single-nucleotide resolution of hearing loss patients, our study can help establish the association between audiometric patterns and methylation status in age-related hearing loss patients.

## INTRODUCTION

Age-related hearing loss (ARHL) or Presbycusis is the most common sensory disorder present in the elderly population [1]. ARHL arises from the inescapable decline in hearing ability that happens with age. Approximately one in three adults over the age of 65 and half of 85-year-olds have hearing loss, showing a consistent decline in hearing [2]. ARHL can also have psychological effects, it may contribute to depression, isolation, and dementia [3]. The main characteristic of ARHL is the inability to understand high-frequency elements of speech, silent consonants and decrease ability in hearing speech in loud environments. ARHL diagnosis is validated with audiometry and technologies such as hearing aids and cochlear implantations are used to alleviate the various symptoms, yet they do not restore hearing to normal [4].

To date there is no distinct cure for ARHL since the underlying causes of this sensory impairment are still unknown. Anatomically ARHL is linked to several auditory structures and divided into different subcategories. Sensory ARHL is caused by the degeneration of the mechanotransducing cochlear inner and outer hair cells. Metabolic ARHL also known as strial ARHL is due to reduced function within the stria vascularis and neural ARHL which shows as deterioration of the auditory nerve [5] [6]. Mixed and Intermediate types of ARHL were added in 1993 as the latest subcategories. Many believe that the majority of individuals will exhibit a mixed pathology of ARHL where changes to the peripheral lesions and the central auditory simultaneously could contribute to the progression of ARHL [7]. ARHL is considered a multifactorial disorder; it involves intrinsic and extrinsic factors. Many factors have been implicated in the development of ARHL: elements such as biological age, gender, environment, mainly genetic predispositions, oxidative stress, mitochondria role in the aging process, cell death apoptosis and necrosis [8]. Additionally, risk factors such as medication-induced ototoxicity, hypertension, diabetes, otitis media infection history, family history of hereditary hearing loss and accumulation of noise exposure have also been linked to ARHL [9]. Currently our understanding of this complex disease is limited, further studies are imperative to complete the urgent demand for therapeutic intervention to improve age-related auditory decline.

Epigenetic changes are dynamics, they can be caused by environmental conditions and age [10]. Epigenetic regulation of gene expression in the ageing ear coupled with environmental exposure might account for age-related changes to hearing ability in ARHL patients. Current studies have shown that epigenetics modifications influence the inner ear [11]. A better understanding of epigenetic mechanisms in hearing impairment can support the development of new treatments for hearing loss. The link between epigenetic mechanisms and hearing loss is well established, however the specific biological mechanism is not well understood [12]. The epigenetic regulation that is known to primarily influence the inner ear involves DNA methylation [13]). There have been several associations between DNA methylation and ARHL. One study has shown that decrease in the expression of Connexin 26 (Cx26), the protein encoded by the *GJB2* gene which forms gap junctions (GJs), may contribute to the development of ARHL. An increase of CpG methylation at the promoter region of the *GJB2* gene has also been found and the hypermethylation of the promoter region has been associated with Cx26 downregulation [14]. *CDH23* hypermethylation has been shown to be involved in ARHL disorder; It has recently been demonstrated that higher CpG sites methylation levels in *CDH23* are likely to be associated with ARHL. Cadherin 23, also called otocadherin, is a member of a super-family of calcium dependent cell surface adhesion protein. Cadherin 23 protein is known to be a part of lateral and stereo ciliary tip links of the inner ear sensory hair, which then controls the hearing process [15].

In this paper we illustrate an approach that can help improve the difficult task of identifying the main cause for ARHL. We hypothesize that ARHL cases could derive from alterations in epigenetic regulations, this would explain why the main causes of this disease cannot be found by simply looking at the DNA sequence. The objective of this study is to identify an association between DNA methylation changes and Age-related hearing loss

## METHODS

### Subjects

This study was approved by the University of Miami institutional review board. Subjects were recruited from adults attending the outpatient clinic of the University of Miami Ear Institute. All patients completed a questionnaire on demographic information including ethnicity and a medical history focusing on the identification of factors with known effects on hearing such as excess noise, ear diseases, ear trauma, radiation exposure of surgery, chronic illness (i.e., diabetes, cardiovascular disease), manifestations of syndromic deafness (i.e., blindness from retinitis pigmentosa, tegumentary and craniofacial anomalies), and family history of hearing loss. At least a three-generation family history was obtained for each subject and the case classified based on the inheritance pattern in autosomal recessive, autosomal dominant mitochondrial DNA, X-linked or sporadic case.

The inclusion criteria for subjects with ARHL consisted of age greater than 40 years and greater than 30 dB HL hearing loss (bone conduction pure tone average [PTA] of frequencies 500, 1000, 2000, and 4000 Hz). Exclusion criteria included 1) previous history of exposure to excess environmental and occupational noise 2) exposure to toxins or drugs with known ototoxic effects; 3) history of temporal bone trauma 4) history of otologic disorders such as Meniere’s disease, autoimmune hearing loss, etc.; 5) average conductive hearing loss greater than 15 dB hearing loss (HL) in one or two ears, measured at 500,100, and 200 Hz; 6) unilateral or significantly symmetric (greater than 25 dB difference in interaural pure-tone average, PTA) hearing loss. [16] [17].

### Audioprofiles

The recruited subjects with ARHL underwent audiological examination according to current clinical standards (ISO 8253-1, 1989). The audiological examination consisted of measurements of air and bone conduction pure-tone thresholds at 500,1000 2000, 4000, and 8000 Hertz. Air-conduction threshold curves were classified into one of five audiometric configurations or audioprofiles based on previously reported guidelines [18] [19].

1. High-frequency gently sloping ;15-29 dB difference between the average of 0.5-1 kHz and the average of 4-8 kHz
2. High frequency steeply sloping: the difference between the average of 0.5-1 kHz and the average of 4-8 kHz us 30 dB or greater
3. Low-frequency ascending; greater than 15 dB difference between 0.5 and 2kHz
4. Mid-Frequency “U” shaped: greater than 15 dB difference between the average of mid-frequency pure-tones (1-2 kHz), and that of the low tones (0.25 to -0.5 kHz) and high tones (4-8 kHz)
5. Flat: Less than 15 dB difference between the average of 0.25-0.5 kHz, the average of 1-2 kHz, and the average of 4-8 kHz.

The audiometric patterns that were more frequent in the cohort study are “High frequency Steeply Slopping” or HFSS (33%),” High frequency Gently Slopping” or HFGS (31%) and “FLAT” (27%) while the other patterns were less prevalent. No statistical significance was found in terms of gender, age, ear side, and PTA values among the audiometric types [16] [17].

### DNA Isolation and Bisulfite conversion

Blood samples were collected, and genomic DNA was extracted from blood using a standard extraction method. DNA concentrations were measured using the Qubit ^®^ dsDNA HS Assay (Invitrogen Carlsbad, CA). For methylation analysis500 ng of Isolated DNA was treated with bisulfite using the Epigentek Bisulflash DNA Modification Kit.

### CANDIDATE Genes Approach

A list of genes involved in Hearing loss was assembled from an intensive review of the literature of sensorineural hearing loss (SNHL), Age-related Hearing loss (ARHL), both nonsyndromic and syndromic hearing loss as well as association and animal models studies.

### Array-Based Methylation Assay

The Illumina MethylationEPIC BeadChip was used as an array-based methylation profiling. The MethylationEPIC BeadChip (Illumina, San Diego, CA, United States) measures over 850,000 CpG loci across diverse set of functionally relevant genomic regions that include promoters, CpG islands (CGI), CGI shores, intergenic CpGs as well as gene body. Approximately, 500 ng of genomic DNA were bisulfite modified using the EZ-96 DNA Methylation Kit (Zymo Research, Irvine, CA, United States) then processed as indicated by Illumina. The BeadChip images were scanned and the data was analyzed using the R software (version 3,2,4). For quality control probes that had a detection p-value <0.01 were selected for all samples. The Illumina assay utilize a pair of probes (a methylated probe and an unmethylated probe) to measure the intensities of the methylated and unmethylated alleles at the interrogated CpG sites. The methylated and unmethylated signal intensities were quantile normalized for each individual probes. Subsequently both beta and M values were estimated. Beta values represent the ratio of the methylated signal intensity to the sum of both unmethylated and methylated signals after background subtraction (Beta values range from 0 to 1, 0 corresponding to a completely unmethylated site, while 1 respectively represents a fully methylated site). M values were obtained, they have been shown to have superior statistical properties such as homoscedasticity: M values are logit transformations of the beta values, which is *M*= log (beta/1-beta). M-values near 0 signify a similar intensity between the methylated probe and the unmethylated probes; corresponding to a CpG site that is half methylated, while positive M-values suggest that more molecules are more methylated than unmethylated, negative M-values indicate the contrary.

## Statistical Analysis

All analyses were performed with the R software. Linear regression was used to determine the association between patients’ hearing scores and blood DNA methylation levels.

### Methylation Specific Polymerase Chain Reaction Validation

Methylation Specific PCR was performed to validate the methylation data at selected loci. Primers sets were designed using MethPrimer to discriminate between methylated CpG sites from unmethylated alleles. The DNA samples were modified by bisulfite conversion using the Bisulflash DNA Modification Kit (Epigentek) according to the manufacturer’s protocol. Optimized PCR reactions were conducted using the GoTaq Green Master Mix (Promega). The PCR conditions were as follows: reaction volume, 25 μl: Primers, 10 pM; template genomic DNA, 100ng; denaturation at 95° C for 5 minutes and at 95° C for 30 s; at 60° C for 30 s and 72° C for 30 s for 35 cycles followed by a 5-minute final extension at 72° C. Primer sequences for the *ESPN* and *TNFRSF25* genes tested are shown in Table 1.

**Table 1.**
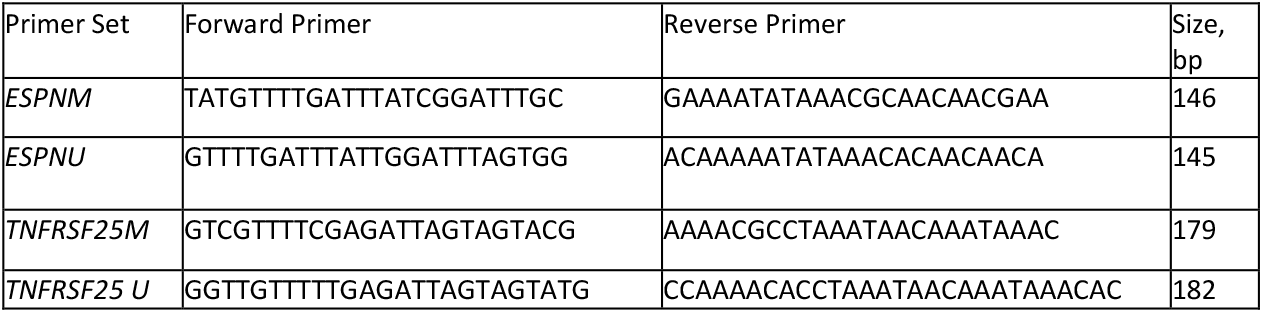
Primers were designed using MethPrimer. The Primer sets used for the amplification were labelled as Methylated (M) and unmethylated (U)

## RESULTS

### Human Subjects

The study consisted of 14 ARHL patients who met the audiometric inclusion criteria as previously described. All findings on audiometric, clinical data were formerly reported [16] [17]. The average age of the subjects was 63 ± 10 years. All subjects were prescreened and found negative for mutations in the *GJB2 g*ene and *GJB6* deletions (GJB6-D13S1830 and GJB6-D13S1854) as well as pathogenic mitochondrial variants (MT-RNR1:111.1555 A>G; MT-TL1: m.3243 A>G). The Illumina MethylationEpic BeadChip microarray was used to perform an epigenome-wide association study.

### Robust Association between CpG sites and Hearing Thresholds

Linear regression analysis was used to assess the relationship between the patients hearing threshold (output variable, x) and the m-values scores (output variable, y). We have determined an association between ARHL patients hearing thresholds and the M-values obtained for the individual CpG sites. A notable finding from our analysis is that two contiguous CpGs located downstream of the *ESPN* gene (cg114044945) and 3’UTR of The Tumor Necrosis Factor Receptor genes *TNFRSF25* (cg2724823) have shown positive correlation with the hearing thresholds (**Figure 1**). Similarly in a sex-stratified manner, this effect is observed at every audiometric frequency (0.5 kHZ-8 kHZ): these 2 genes show statistical evidence of an association between the patient hearing level and the level of CpG sites methylation in females (**Figure 2**) and males (**Figure 3**). These two adjoining CpG cites increase methylation as patients hearing aggravates. These CpGs are located in an area identified as a distal enhancer by the ENCODE Registry of Candidate cis-Regulatory Elements (cCREs). Further, they are positioned in a CpG island and are in close proximity to expression quantitative trait locus (eQTL) for both ESPN and *TNFRSF25* genes identified in GTEx (**Additional File 1**). QQ plots and Manhattan plots were generated using the qqman package in R. QQ plots showed inflation, likely due differences in population structure that are not fully captured by the principal components, as the samples herein have some Hispanic and African American ancestry (**Additional File 1 and 2**). Manhattan plots demonstrating association between hearing scores and DNA methylation of ARHL patients for low and high frequencies were shown. The P values are represented in genomic order by chromosomes and positions on the chromosome (x-axis). All CpGs labeled in the Manhattan plots show nominal association (p < 1E-05) with the ARHL patients hearing threshold at each audiometric frequency (0.5 kHZ - 8 kHZ) but do not reach genome-wide association (p < 5E-08) (**Figure 4 and Figure 5**). The list of nominally significant CpGs (p < 1E-05) sorted by p-value for low and high audiometric frequencies are shown in (**Additional File 3: Table 1a**,**1b**,**1c**,**1d**,**1e**,**1f**).

**Fig. 1.**
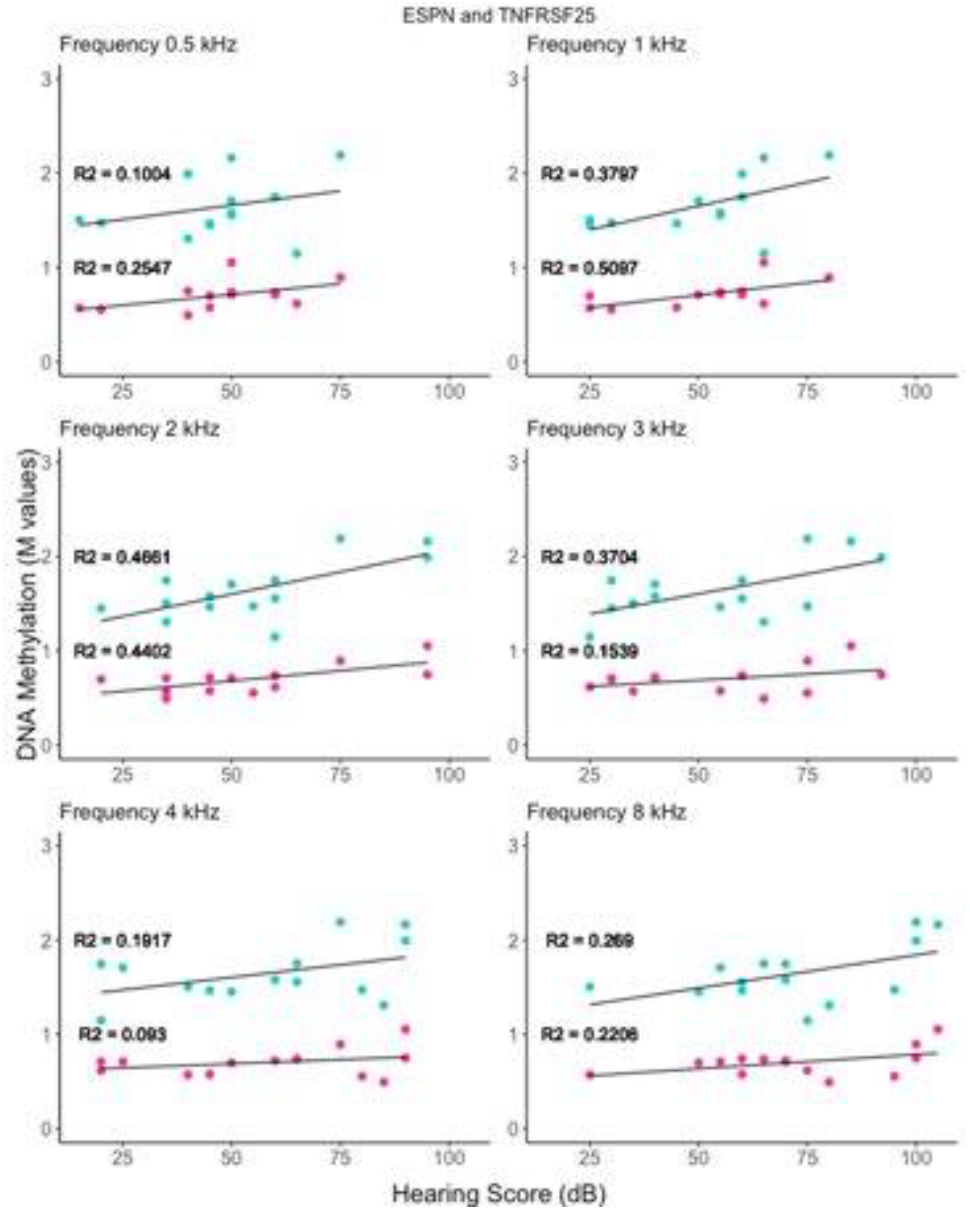
Two Adjacent CpG sites, cg11404945 and cg27224823 located between ESPN and TNFRSF25, show an increase in methylation levels as the patients hearing deteriorate. A positive correlation between the hearing level and the level of CpG island Methylation at each frequency, shown here in the linear regression analysis performed. The CpG cite cg11404945 is located in the downstream region of ESPN, position 6521138 (hg19)on chromosome 1. Mutation in ESPN can cause DFNB36, nonsyndromic dominant hearing loss and USH1M. The CpG cite cg27224823, corresponds to the TNFRSF25 genes, is located on chromosome 1, position 6521268 (hg19). TNFRSF25 Play vital role in regulating cell proliferation, differentiation, and apoptosis

**Fig 2.**
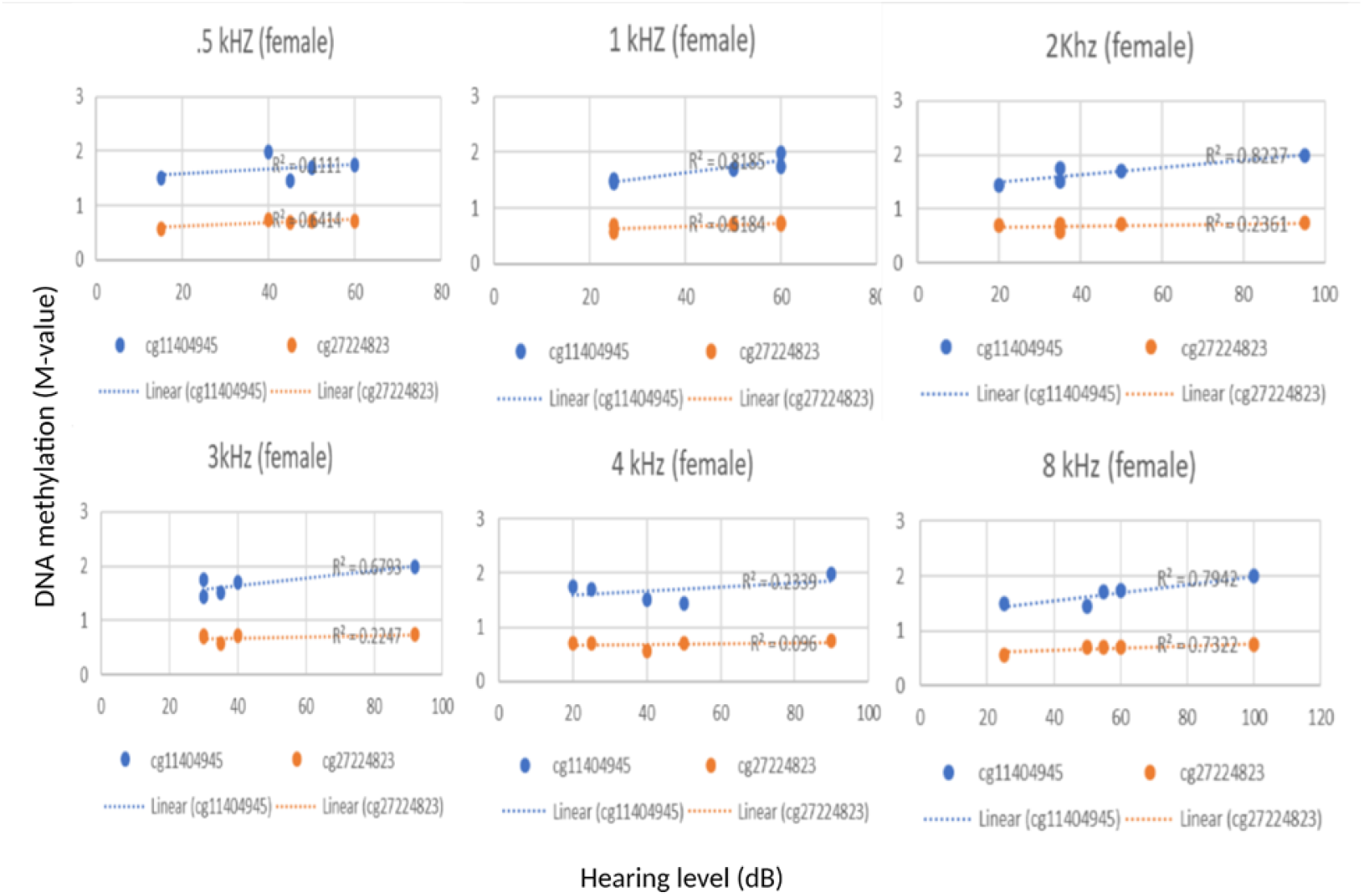
In a sex-stratified manner, in all frequency, a clear correlation between the hearing measurements and the level of methylation in the ESPN and Tumor Necrosis Factor Genes has been observed in females.

**Fig 3.**
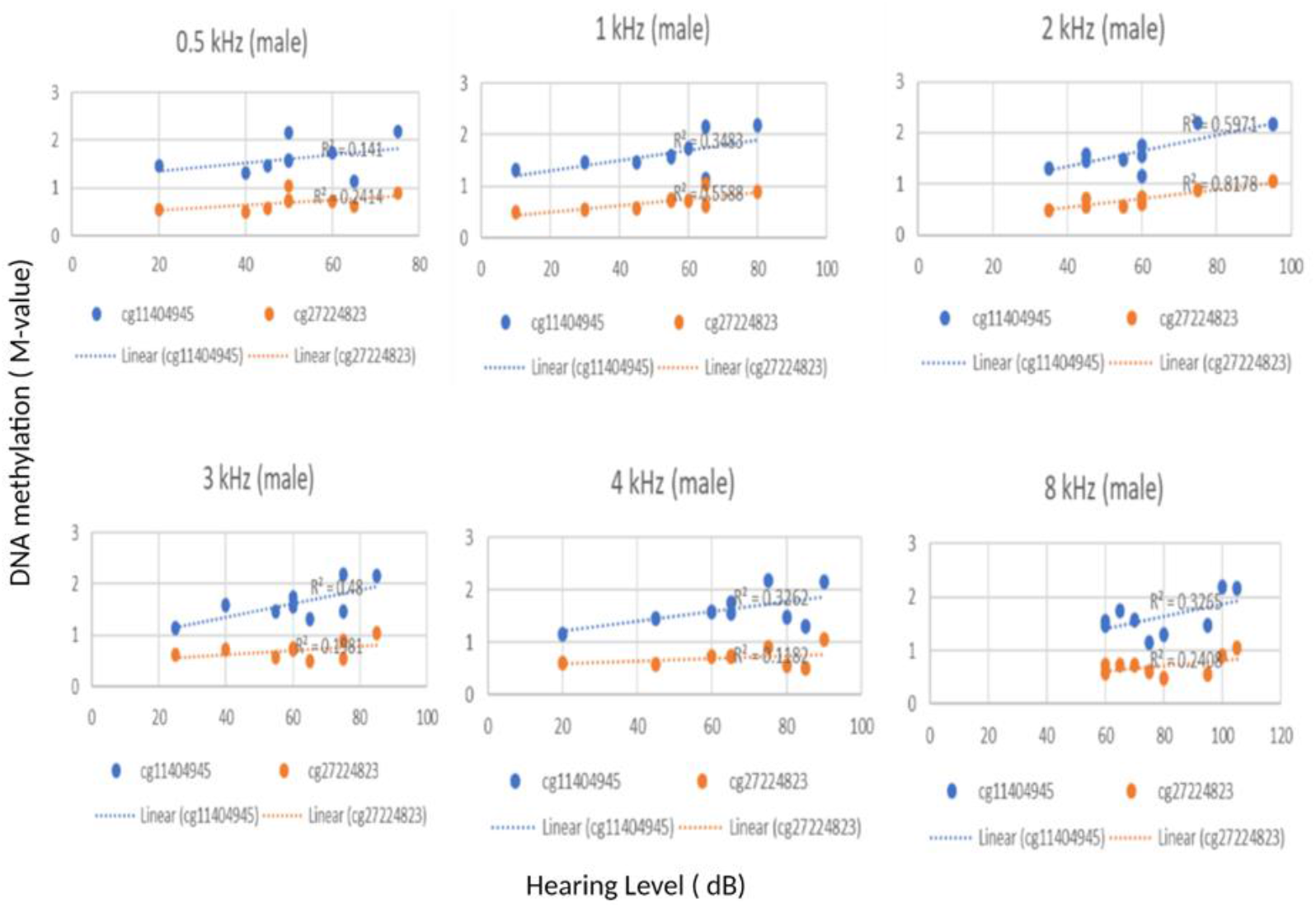
In as sex-stratified approach, a positive correlation can be observed in all the male patients from our cohort, as the patients hearing scores aggravates, an increase in methylation of the *ESPN* and *TNFRSF25* CpG cites methylation level increase.

**Fig 4:**
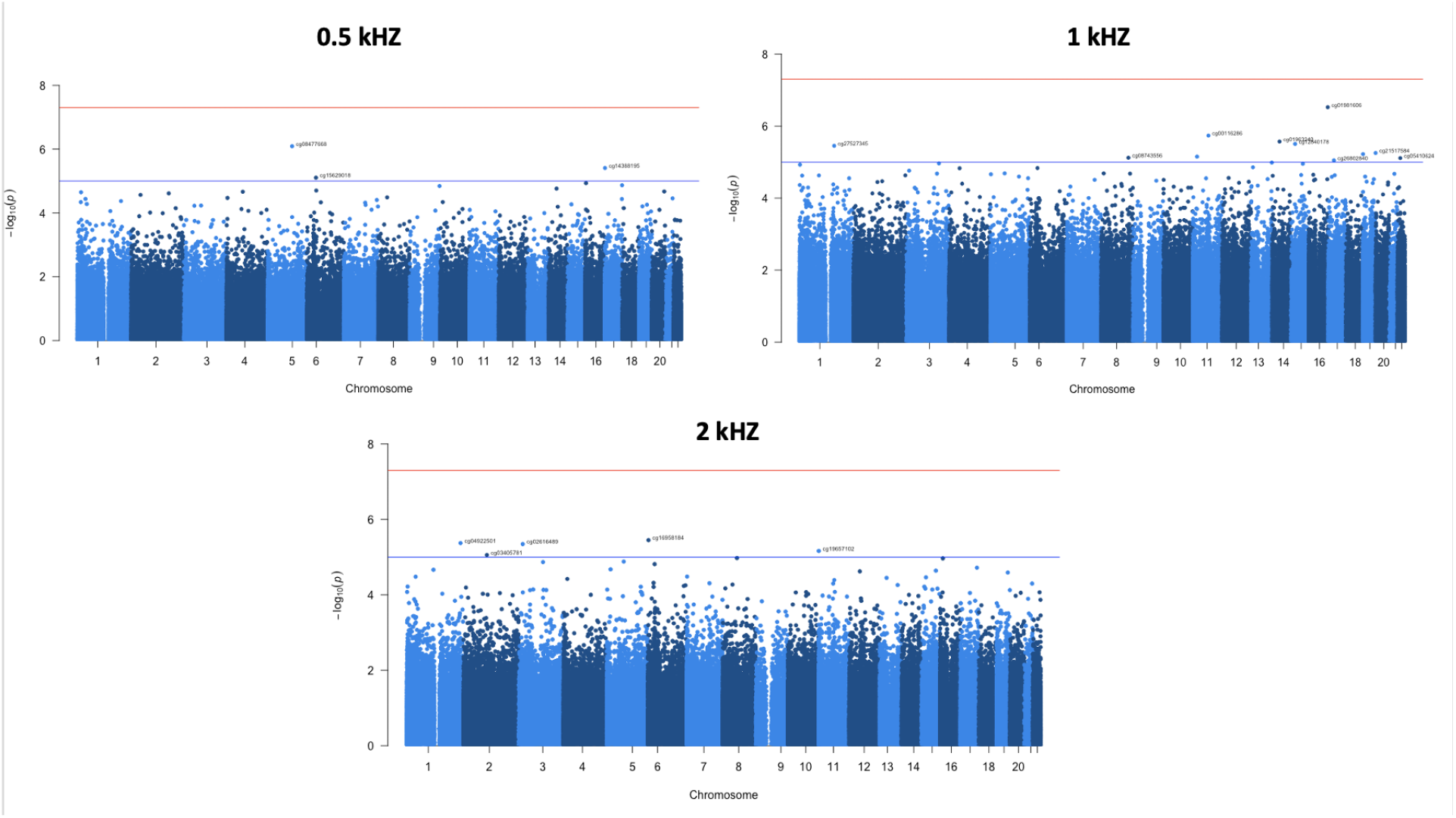
Manhattan plot of the genome-wide DNA methylation analysis in presbycusis patients Each plot depicts the - log 10 p-value of the association between CpG sites and hearing threshold at various low audiometric frequencies. The Blue line is drawn to separate the CpG cites that surpassed a nominal significance threshold of p-values 1×10^−5^. The red line is drawn to separate the CpG cites that surpassed genome-wide significance threshold of p-values < 5×10^−8^. However, none of the CpGs reached genome-wide significance.

**Fig 5:**
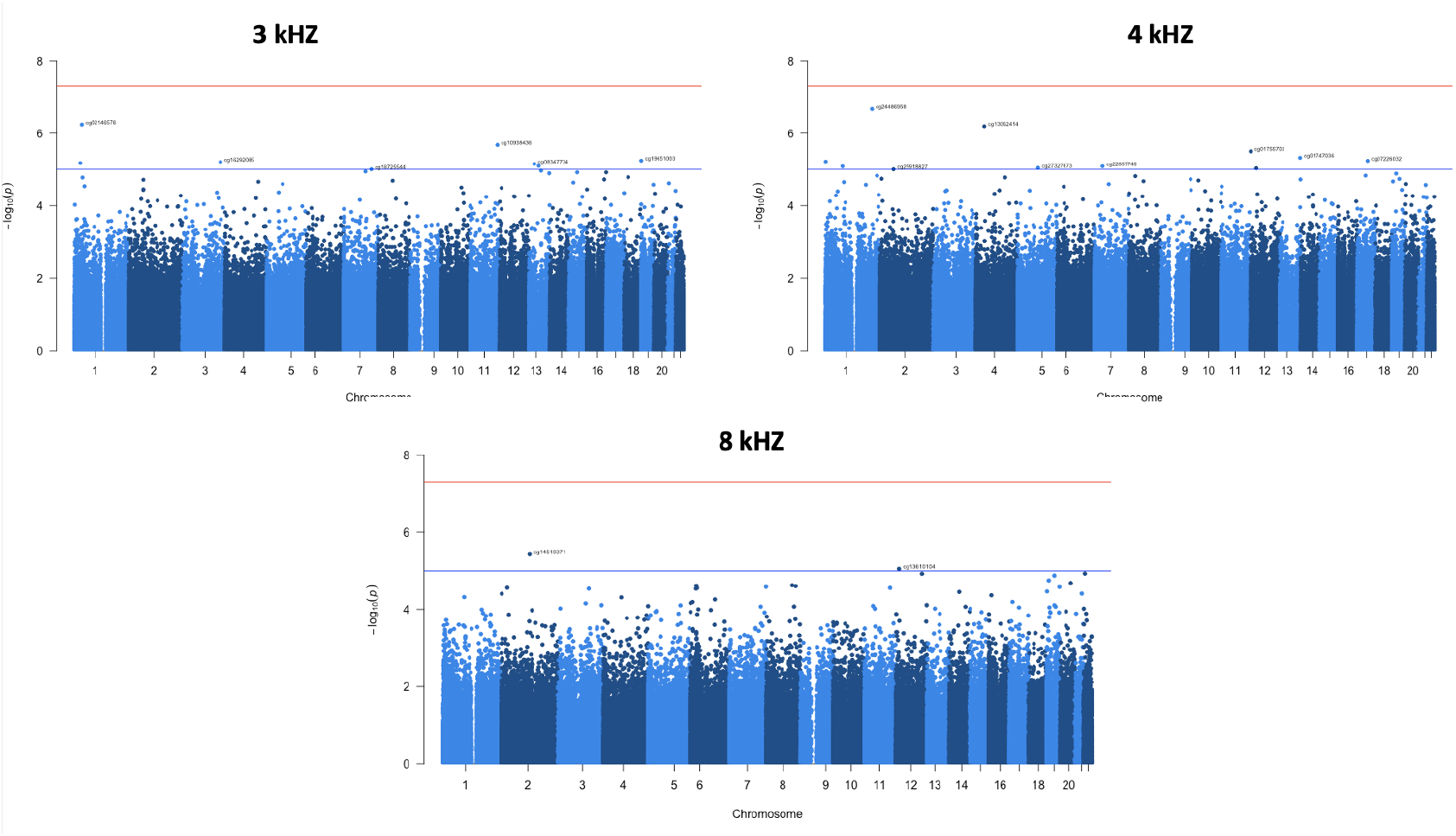
Manhattan Plot depicting the -log _10_ p-value of the association between CpG sites and hearing threshold at high audiometric frequencies. The *P* values are represented in genomic order by chromosomes and positions on the chromosome (x-axis). The values on the y-axis correspond to the -log_10_ of the *P* values. The blue line is drawn to separate the CpG cites that surpassed a nominal significance threshold of p-values < 1×10^−5^. The red line is drawn to separate the CpG cites that surpassed genome-wide significance threshold of p-values < 5×10^−8^. However, none of the CpGs reached genome-wide significance.

### Correlation between Patients DNA Methylation level and Hearing Score at 8 kHZ

ARHL is known to be typified in high frequencies. In this study we proceeded to divide our subjects’ patients based on hearing audioprofiles at 8 kHZ. Group one comprised patients with mild hearing loss (26 dB – 40 dB), the second group was made up of patients with moderate hearing loss (41 dB-70 dB) while group three included patients with severe hearing loss (90+ dB) (**Figure 6**). Our data shows higher level of methylation in the *ESPN* and *TNFRSF25* CpG sites in patients with severe hearing loss at 8 KZ compared to the patients with mild and moderate hearing loss.

**Fig 6:**
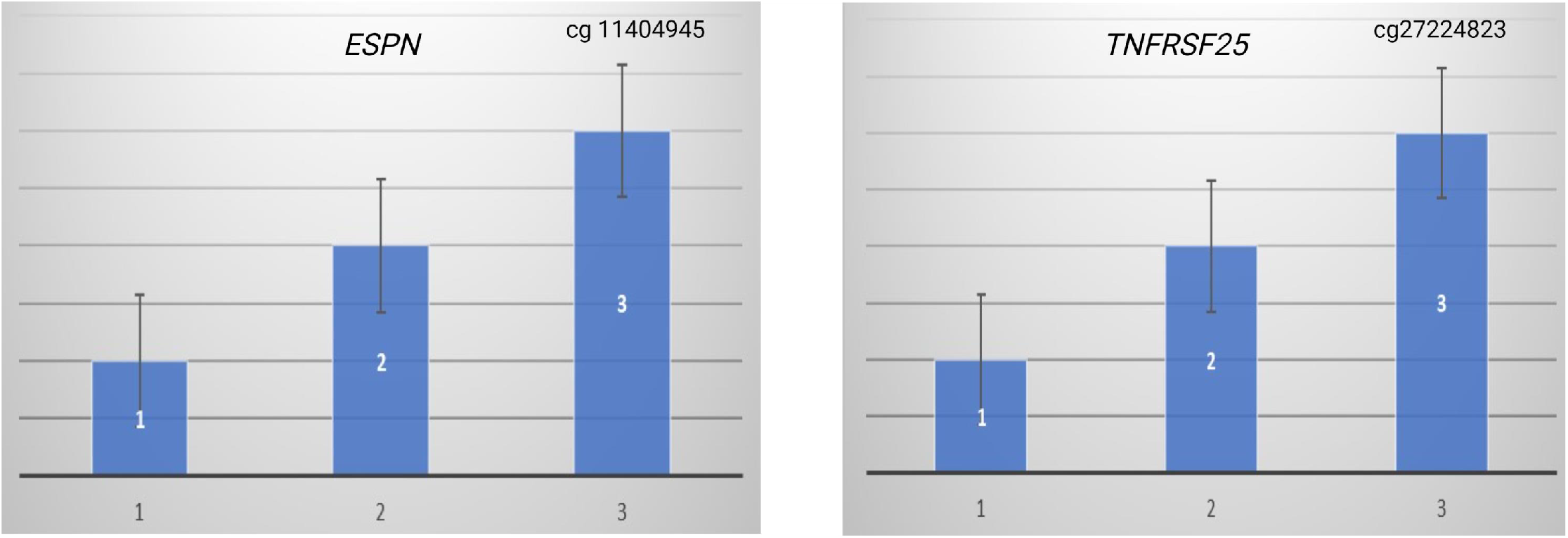
At the high audiometric frequency (8 KHz), the group 3 comprised of patients with severe hearing loss showed higher methylation level in the ESPN and TNFRSFR25 CpG sit0065s compared to the methylation level in the group containing moderate hearing loss (group 2) and mild/normal patients (group 1).

### Methylation Specific PCR Validation

Validation of selective methylated gene-specific CpGs identified from the Illumina MethylationEpic BeadChip was performed using Methylation-Specific PCR of the ARHL patients’ genomic DNA. We showed the methylation status of the identified CpG cites were consistent with the Methylation Epic microarray (Illumina) assay, which interrogates the DNA methylation of 865,000 CpG sites. Our results thus validating the methylation status of the *ESPN* (cg11404945) and *TNFRFS25* (cg27224823) gene. (**Figure 7**). Therefore, the methylation status of the *ESPN* and *TNFRSF25* genes suggest the possible utilization of these hearing-loss subjective genes as epigenetic signatures, hence proving that DNA methylation can be used as a tool for epigenetic-based therapeutic strategies.

**Fig 7:**
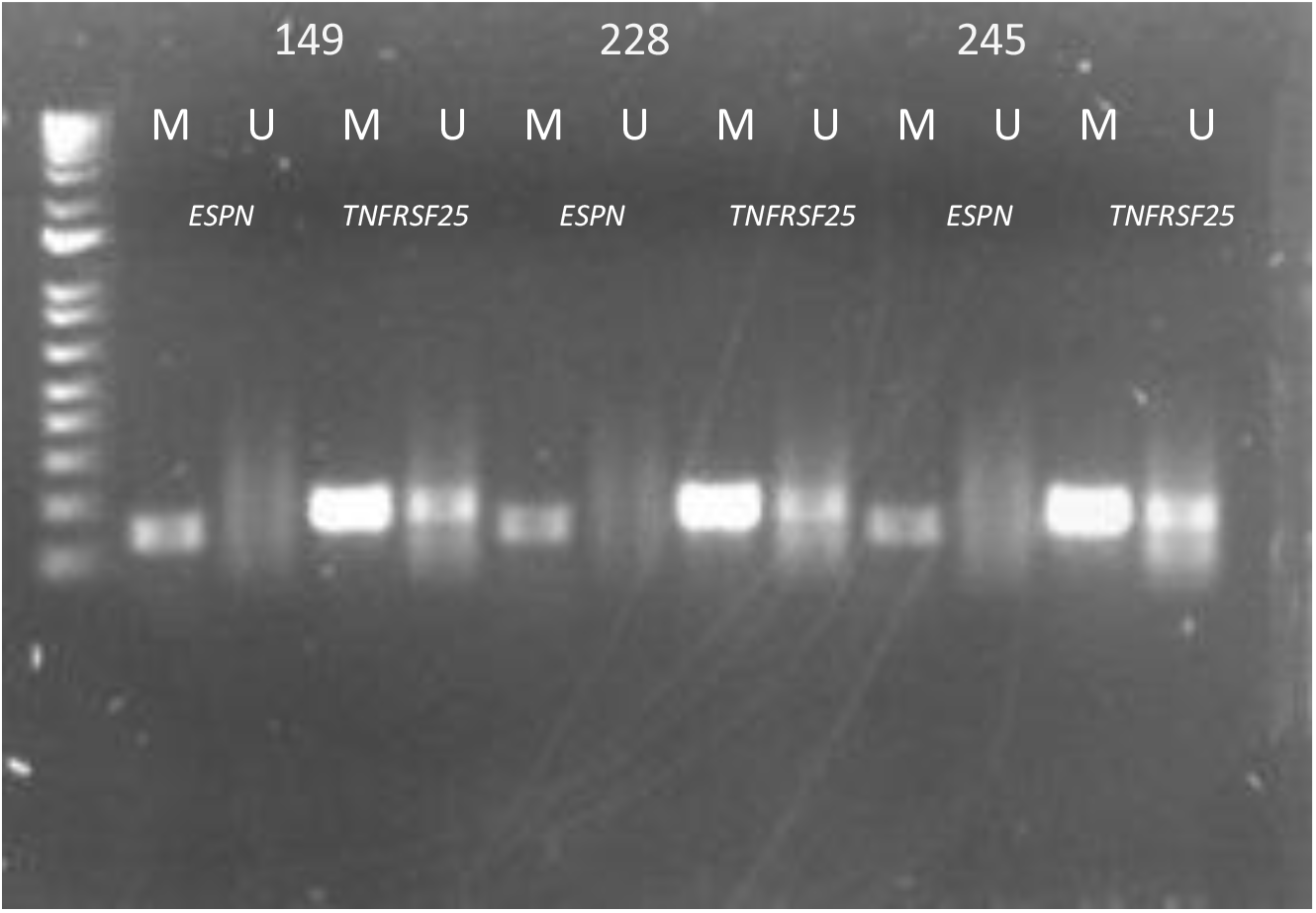
The MSP Primer sets used for amplification are designated as unmethylated (U), methylated (M). Amplification of bisulfite-treated DNA from Presbycusis patients denoted here as #149,228,245 was performed for all DNA samples. A) The PRC products are depicted above using both sets of primer A) *ESPN*, B) *TNFRSF25* genes, validating the methylation values obtained from the Illumina Microarray.

## DISCUSSION

In this study, we performed an epigenome-wide association method, we have identified novel CpG sites that can be associated with ARHL. Methylation of DNA CpG sites is associated with disease state, ageing and environmental factors [9]. Given the polygenic and multifactorial nature of ARHL, with this study we inquired into the extent of involvement of epigenetic alteration in this disorder.

Numerous studies have verified the contribution of epigenetics to various disorders including hearing loss [13]. DNA methylation has been found to play a role in the inner ear, a dynamic pattern of DNA methylation has been observed during key time points in the development of the inner ear [22]. DNA methylation is catalyzed by three important methyltransferases: DNMT1, DNMT3A and DNMT3B. These methyltransferases are positioned on the short arm of chromosome 19 in bands 19p13.3p13.29. Each of these methyltransferases have different functions in the methylation process [23]. Furthermore, epigenetic mechanism has been linked to the development of the ear including DNA methylation; DNMT3 has been shown to be directly correlated to poor physiological embryonic development which in turn can result in hearing loss [24]. The role of epigenetic in deafness is just commencing to be understood however the epigenetics field has seen several developments in uncovering biological mechanisms of gene expression and has propelled the development of epigenetic therapeutics [25].

In this present report a strong correlation between elevated CpG in two contiguous sites located in the intergenic region between the genes *ESPN* and *TNFRSF25*. These CpG sites localize in a CpG island, in a region predicted to act as a transcriptional enhancer and that contains eQTLs for *ESPN* and *TNFRSF25*. This suggest that the altered methylation at these sites could affect expression of the *ESPN* and *TNFRSF25* genes. Espins are actin bundling proteins found in hair cells stereocilia, mutation of the *ESPN* gene can cause hearing loss and vestibular dysfunction in the *jerker* mouse [26]. Additionally, an in-frame deletion of human *ESPN* has been associated with USH1M [27]. The TNFRSF25 (TNF Receptor Superfamily Number 25) with an ability to bind necrosis factors via an extracellular cysteine-rich domain, and has been shown to play a role in the death receptor signaling pathway and apoptosis [28]. Using a sex-stratified strategy, we have also determined in all frequency that there is a positive correlation between our patients’ hearing measurement and the level of methylation in the *ESPN* and T*NFRSF25* as well. Future comprehensive studies are warranted to test the concord of the *ESPN* and T*NFRSF25* methylation and gene transcription levels. Our results demonstrate that the epigenetic field, complimentary to our current knowledge of the gene networks can enhance our understanding of the genome in response to intracellular and environmental factors.

There are several limitations to our study that should be recognized. First the limited samples size of ARHL patients helped us in achieving an association study, future studies will be conducted by adding age-gender-matched controls samples from the same geographical areas as that of the subject. A second limitation is the usage of DNA samples from ARHL patients’ peripheral blood due to the inaccessibility to inner ear tissues. In conclusion our study highlights the implication of DNA methylation; a main epigenetic marker in regulating gene transcriptional regulation and preserving genome stability association with ARHL. These results suggest that epigenetic mechanisms can be a contributor to hearing loss during the ageing process.

## Supporting information

The list of nominally significant CpGs (p < 1E-05) sorted by p-value for low and high audiometric frequencies are shown in (Additional File 3: Ta

## Data Availability

Availability of data and material
The datasets generated and/or analyzed during the present study are available from the corresponding author on reasonable requests

## Declarations

### Ethics approval and consent to participate

This study was approved by the University of Miami institutional Review Board. DNA samples included in this study were collected from adults subjects attending the outpatient clinic of the University of Miami Ear Institute. All patients provided written informed consent.

### Consent for publication

All participants provided written informed consent for publication.

### Availability of data and material

The datasets generated and/or analyzed during the present study are available from the corresponding author on reasonable requests.

### Authors Contributions

M.V.R., D.Y., S.B., F.G., X.L contributed to methodology, conceptualization, writing of the original draft. D.G, J.Y., N.H., PT contributed to the data acquisition and interpretation, and content revision.

## Acknowledgements

We would like to thank the National Institute of Health/National Institute on Deafness and Other Communication Disorders for the support.

## Figure Legends

**Additional File 1:** UCSC genome browser tracks. Top track (custom): showing the location of the CpGs that associate with hearing score (cg114044945 and cg2724823, orange bars) and the location of the eQTL variants as identified by GTEx (yellow bars, https://gtexportal.org/). Second track: Gencode genes. Third track: ENCODE regulatory elements (cCREs). Bottom track: CpG Islands. The rs2986758 is an eQTL for both ESPN and TNFRSF25, whereas rs 147172610 and 54481403 are eQTLs for ESPN.

**Additional File 1 and 2**: QQ plots and Manhattan plots were generated using the qqman package in R.

**Additional File 3**: List of nominally significant CpGs (p < 1E-05) sorted by p-value for low and high audiometric frequencies.

**Supplementary:**
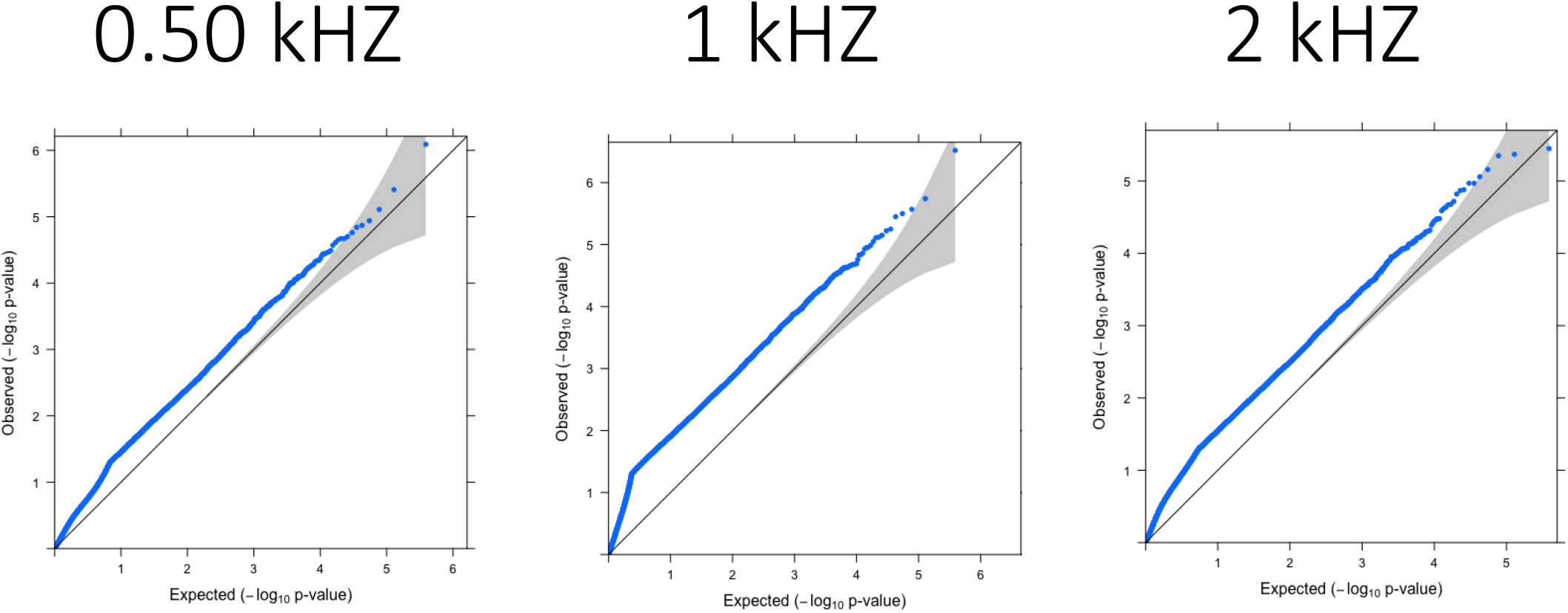
QQ plots – low freq

**Supplementary:**
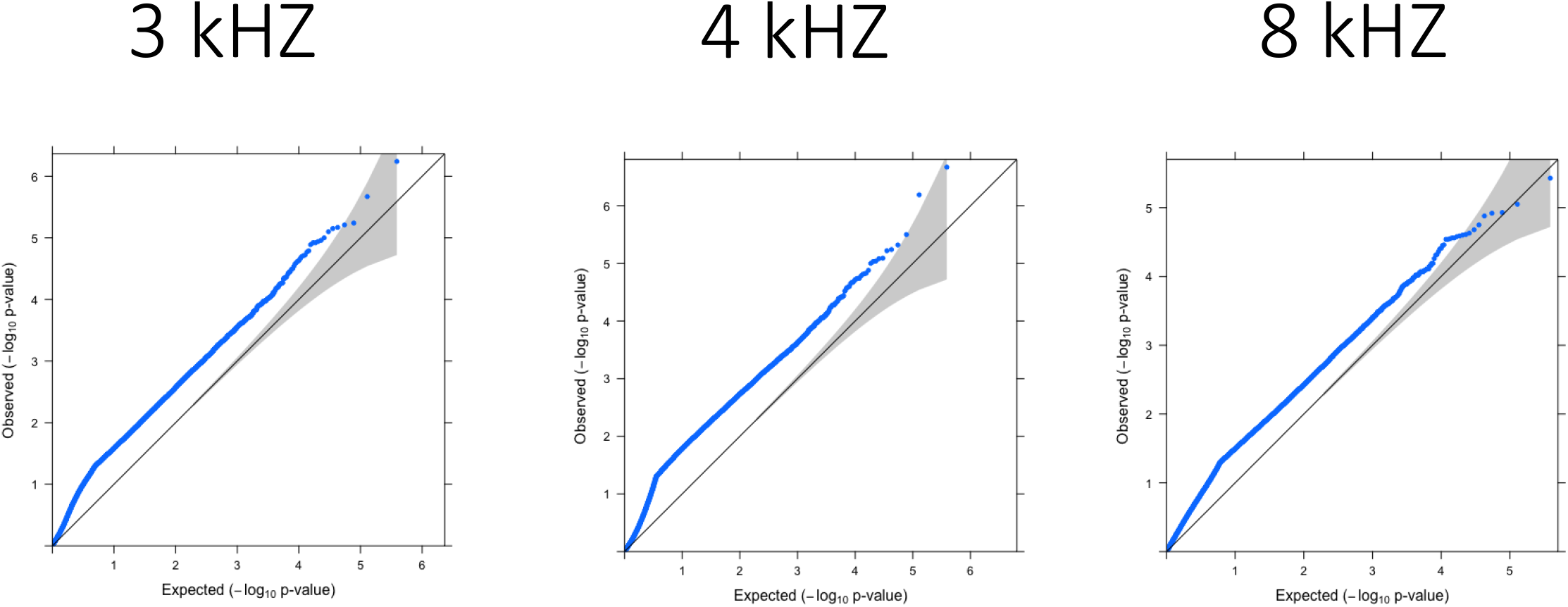
QQ plots – high freq

**Supplementary figure X:**
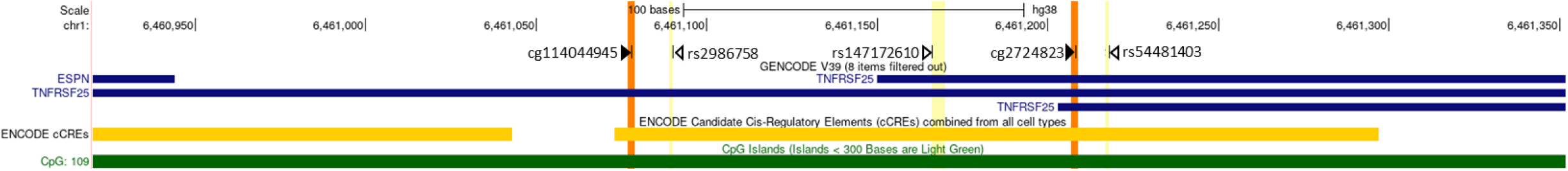
UCSC genome browser tracks. Top track (custom): showing the location of the CpGs that associate with hearing score (cg114044945 and cg2724823, orange bars) and the location of the eQTL variants as identified by GTEx (yellow bars, https://gtexportal.org/). Second track: Gencode genes. Third track: ENCODE regulatory elements (cCREs). Bottom track: CpG Islands. The rs2986758 is an eQTL for both *ESPN* and *TNFRSF25*, whereas rs 147172610 and rs 54481403 are eQTLs for *ESPN*.

## Bibliography

1. Ashapkin, V. V., L. I. Kutueva and B. F. Vanyushin (2019). “Epigenetic Clock: Just a Convenient Marker or an Active Driver of Aging?” Adv Exp Med Biol 1178: 175–206.

2. Bouzid, A., I. Smeti, A. Chakroun, S. Loukil, A. A. Gibriel, M. Grati, A. Ghorbel and S. Masmoudi (2018). “CDH23 Methylation Status and Presbycusis Risk in Elderly Women.” Front Aging Neurosci 10: 241.

3. Bowl, M. R. and S. J. Dawson (2019). “Age-Related Hearing Loss.” Cold Spring Harb Perspect Med 9(8).

4. Cheslock, M. and O. De Jesus (2021). Presbycusis. StatPearls. Treasure Island (FL), StatPearls Publishing Copyright © 2021, StatPearls Publishing LLC.

5. Du, P., X. Zhang, C. C. Huang, N. Jafari, W. A. Kibbe, L. Hou and S. M. Lin (2010). “Comparison of Betavalue and M-value methods for quantifying methylation levels by microarray analysis.” BMC Bioinformatics 11: 587.

6. Franklin, T. B. and I. M. Mansuy (2010). “Epigenetic inheritance in mammals: evidence for the impact of adverse environmental effects.” Neurobiol Dis 39(1): 61–65.

7. Gates, G. A. and J. H. Mills (2005). “Presbycusis.” Lancet 366(9491): 1111–1120.

8. Maksimovic, J., B. Phipson and A. Oshlack (2016). “A cross-package Bioconductor workflow for analysing methylation array data.” F1000Res 5: 1281.

9. Mittal, R., N. Bencie, G. Liu, N. Eshraghi, E. Nisenbaum, S. H. Blanton, D. Yan, J. Mittal, C. T. Dinh, J. I. Young, F. Gong and X. Z. Liu (2020). “Recent advancements in understanding the role of epigenetics in the auditory system.” Gene 761: 144996.

10. Nolan, L. S. (2020). “Age-related hearing loss: Why we need to think about sex as a biological variable.” J Neurosci Res 98(9): 1705–1720.

11. Ohlemiller, K. K. (2004). “Age-related hearing loss: the status of Schuknecht’s typology.” Curr Opin Otolaryngol Head Neck Surg 12(5): 439–443.

12. Singmann, P., D. Shem-Tov, S. Wahl, H. Grallert, G. Fiorito, S. Y. Shin, K. Schramm, P. Wolf, S. Kunze, Y. Baran, S. Guarrera, P. Vineis, V. Krogh, S. Panico, R. Tumino, A. Kretschmer, C. Gieger, A. Peters, H. Prokisch, C. L. Relton, G. Matullo, T. Illig, M. Waldenberger and E. Halperin (2015). “Characterization of whole-genome autosomal differences of DNA methylation between men and women.” Epigenetics Chromatin 8: 43.

13. Tu, N. C. and R. A. Friedman (2018). “Age-related hearing loss: Unraveling the pieces.” Laryngoscope Investig Otolaryngol 3(2): 68–72.

14. Wang, J. and J. L. Puel (2020). “Presbycusis: An Update on Cochlear Mechanisms and Therapies.” J Clin Med 9(1).

15. Wu, X., Y. Wang, Y. Sun, S. Chen, S. Zhang, L. Shen, X. Huang, X. Lin and W. Kong (2014). “Reduced expression of Connexin26 and its DNA promoter hypermethylation in the inner ear of mimetic aging rats induced by d-galactose.” Biochem Biophys Res Commun 452(3): 340–346.

16. Yamasoba, T., F. R. Lin, S. Someya, A. Kashio, T. Sakamoto and K. Kondo (2013). “Current concepts in age-related hearing loss: epidemiology and mechanistic pathways.” Hear Res 303: 30–38.

17. Yang, C. H., T. Schrepfer and J. Schacht (2015). “Age-related hearing impairment and the triad of acquired hearing loss.” Front Cell Neurosci 9: 276.

18. Zhang, L., J. I. Young, L. Gomez, T. C. Silva, M. A. Schmidt, J. Cai, X. Chen, E. R. Martin and L. Wang (2021). “Sex-specific DNA methylation differences in Alzheimer’s disease pathology.” Acta Neuropathol Commun 9(1): 77.

19. Maksimovic, J., B. Phipson, and A. Oshlack, A cross-package Bioconductor workflow for analysing methylation array data. F1000Res, 2016. 5: p. 1281.

20. Du, P., et al., Comparison of Beta-value and M-value methods for quantifying methylation levels by microarray analysis. BMC Bioinformatics, 2010. 11: p. 587.

1. Tu, N.C. and R.A. Friedman, Age-related hearing loss: Unraveling the pieces. Laryngoscope Investig Otolaryngol, 2018. 3(2): p. 68–72.

2. Wang, J. and J.L. Puel, Presbycusis: An Update on Cochlear Mechanisms and Therapies. J Clin Med, 2020. 9(1).

3. Gates, G.A. and J.H. Mills, Presbycusis. Lancet, 2005. 366(9491): p. 1111–20.

4. Cheslock, M. and O. De Jesus, Presbycusis, in StatPearls. 2021, StatPearls Publishing Copyright © 2021, StatPearls Publishing LLC.: Treasure Island (FL).

5. Schuknecht, H.F., Presbycusis. Laryngoscope, 1955. 65(6): p. 402–19.

6. Ohlemiller, K.K., Age-related hearing loss: the status of Schuknecht’s typology. Curr Opin Otolaryngol Head Neck Surg, 2004. 12(5): p. 439–43.

7. Bowl, M.R. and S.J. Dawson, Age-Related Hearing Loss. Cold Spring Harb Perspect Med, 2019. 9(8).

8. Yang, C.H., T. Schrepfer, and J. Schacht, Age-related hearing impairment and the triad of acquired hearing loss. Front Cell Neurosci, 2015. 9: p. 276.

9. Yamasoba, T., et al., Current concepts in age-related hearing loss: epidemiology and mechanistic pathways. Hear Res, 2013. 303: p. 30–8.

10. Kang, J.G., et al., Regulation of gene expression by altered promoter methylation using a CRISPR/Cas9-mediated epigenetic editing system. Sci Rep, 2019. 9(1): p. 11960.

11. Layman, W.S. and J. Zuo, Epigenetic regulation in the inner ear and its potential roles in development, protection, and regeneration. Front Cell Neurosci, 2014. 8: p. 446.

12. Provenzano, M.J. and F.E. Domann, A role for epigenetics in hearing: Establishment and maintenance of auditory specific gene expression patterns. Hear Res, 2007. 233(1-2): p. 1–13.

13. Mittal, R., et al., Recent advancements in understanding the role of epigenetics in the auditory system. Gene, 2020. 761: p. 144996.

14. Wu, X., et al., Reduced expression of Connexin26 and its DNA promoter hypermethylation in the inner ear of mimetic aging rats induced by d-galactose. Biochem Biophys Res Commun, 2014. 452(3): p. 340–6.

15. Bouzid, A., et al., CDH23 Methylation Status and Presbycusis Risk in Elderly Women. Front Aging Neurosci, 2018. 10: p. 241.

16. Angeli, S.I., et al., Audioprofiles and antioxidant enzyme genotypes in presbycusis. Laryngoscope, 2012. 122(11): p. 2539–42.

17. Bared, A., et al., Antioxidant enzymes, presbycusis, and ethnic variability. Otolaryngol Head Neck Surg, 2010. 143(2): p. 263–8.

18. Demeester, K., et al., Audiometric shape and presbycusis. Int J Audiol, 2009. 48(4): p. 222–32.

19. Liu, X. and L. Xu, Nonsyndromic hearing loss: an analysis of audiograms. Ann Otol Rhinol Laryngol, 1994. 103(6): p. 428–33.

20. Maksimovic, J., B. Phipson, and A. Oshlack, A cross-package Bioconductor workflow for analysing methylation array data. F1000Res, 2016. 5: p. 1281.

21. Du, P., et al., Comparison of Beta-value and M-value methods for quantifying methylation levels by microarray analysis. BMC Bioinformatics, 2010. 11: p. 587.

22. Yizhar-Barnea, O., et al., DNA methylation dynamics during embryonic development and postnatal maturation of the mouse auditory sensory epithelium. Sci Rep, 2018. 8(1): p. 17348.

23. Edwards, J.R., et al., DNA methylation and DNA methyltransferases. Epigenetics Chromatin, 2017. 10: p. 23.

24. Li, Q., P.J. Hermanson, and N.M. Springer, Detection of DNA Methylation by Whole-Genome Bisulfite Sequencing. Methods Mol Biol, 2018. 1676: p. 185–196.

25. García-Giménez, J.L., et al., Epigenetic biomarkers: A new perspective in laboratory diagnostics. Clin Chim Acta, 2012. 413(19-20): p. 1576–82.

26. Donaudy, F., et al., Espin gene (ESPN) mutations associated with autosomal dominant hearing loss cause defects in microvillar elongation or organisation. J Med Genet, 2006. 43(2): p. 157–61.

27. Ahmed, Z.M., et al., Inframe deletion of human ESPN is associated with deafness, vestibulopathy and vision impairment. J Med Genet, 2018. 55(7): p. 479–488.

28. Schreiber, T.H., D. Wolf, and E.R. Podack, The role of TNFRSF25:TNFSF15 in disease… and health? Adv Exp Med Biol, 2011. 691: p. 289–98.

