## Supplementary material for "DNA Methylation Study in Presbycusis Patients": The list of nominally significant CpGs (p < 1E-05) sorted by p-value for low and high audiometric frequencies are shown in (Additional File 3: Ta

Frequency 0.50 kHZ

**Table 1a:** Top 10 CpG at audiometric frequency 0.5 kHZ , sorted by p- value.

|  | CpG | Chr | Pos | Gene | Estimate | SE | p-value |
| --- | --- | --- | --- | --- | --- | --- | --- |
| 1 | cg24693520 | 21 | 38445266 | TTC3; PIGP | 26.16 | 6.20 | 0.00177 |
| 2 | cg07241909 | 11 | 17566163 | USH1C | -17.59 | 4.50 | 0.00291 |
| 3 | cg12573502 | 11 | 2485281 | KCNQ1 | -79.89 | 22.56 | 0.00534 |
| 4 | cg25408700 | 21 | 43726165 | TMPRSS3 | -80.27 | 23.54 | 0.00665 |
| 5 | cg26857837 | 17 | 79477974 | ACTG1 | 41.68 | 12.76 | 0.00849 |
| 6 | cg07449450 | 11 | 17566051 | USH1C | -13.59 | 4.34 | 0.0106 |
| 7 | cg22803950 | 21 | 43109630 | NCRNA00111 | 52.46 | 16.80 | 0.0108 |
| 8 | cg13817341 | 16 | 89977788 | TCF25 | -65.08 | 20.92 | 0.0110 |
| 9 | cg16766640 | 11 | 14277324 | SPON1 | -98.32 | 31.86 | 0.0115 |
| 10 | cg10698098 | 6 | 160148397 | WTAP; SOD2 | -40.34 | 13.61 | 0.0141 |

Frequency 1 kHZ

**Table 1b:** Top 10 CpG at audiometric frequency 1 kHZ , sorted by p- value.

|  | CpG | Chr | Pos | Gene | Estimate | SE | p |
| --- | --- | --- | --- | --- | --- | --- | --- |
| 1 | cg11288641 | chr21 | 42688618 | FAM3B | -45.36 | 8.88 | 0.000458 |
| 2 | cg00800038 | chr16 | 89945340 | TCF25 | -76.84 | 16.01 | 0.000725 |
| 3 | cg05126444 | chr21 | 38629859 | DSCR3 | -49.55 | 11.40 | 0.00144 |
| 4 | cg23919534 | chr1 | 6507804 | ESPN | 66.02 | 15.58 | 0.00172 |
| 5 | cg04973239 | chr21 | 40714455 | HMGN1 | -132.3 | 31.863 | 0.00197 |
| 6 | cg21130221 | chr11 | 2848310 | KCNQ1 | -47.66 | 11.97 | 0.00259 |
| 7 | cg27224823 | chr1 | 6521268 | TNFRSF25 | 106.84 | 27.55 | 0.00307 |
| 8 | cg07456546 | chr21 | 43726315 | TMPRSS3 | 59.23 | 15.34 | 0.00315 |
| 9 | cg19767477 | chr5 | 127420684 | SLC12A2 | -46.80 | 12.167 | 0.00323 |
| 10 | cg16615040 | chr21 | 41516861 | DSCAM; TMPRSS3 | 87.67 | 22.82 | 0.00325 |

Frequency 2 kHZ

**Table 1c:** Top 10 CpG sites at audiometric frequency 2 kHZ , sorted by p- value.

|  | CpG | Chr | Pos | Gene | Estimate | SE | p-value |
| --- | --- | --- | --- | --- | --- | --- | --- |
| 1 | cg19767477 | 5 | 127420684 | SLC12A2 | -58.12 | 9.90 | 0.000157 |
| 2 | cg14527262 | 4 | 1202653 | SPON2 | 49.23 | 10.35 | 0.000772 |
| 3 | cg14696348 | 16 | 89984517 | MC1R | 168.3 | 35.85 | 0.000847 |
| 4 | cg23049234 | 11 | 2595718 | KCNQ1 | 139.51 | 34.46 | 0.00232 |
| 5 | cg03660952 | 11 | 2644418 | KCNQ1 | 279.5 | 69.21 | 0.00236 |
| 6 | cg03215005 | 21 | 42792304 | MX1 | 41.40 | 10.55 | 0.00285 |
| 7 | cg21505925 | 21 | 38807423 | DYRK1A | -35.19 | 9.80 | 0.00492 |
| 8 | cg18217706 | 21 | 43734404 | TFF3 | -113.1 | 32.33 | 0.00575 |
| 9 | cg11404945 | 1 | 6521138 | ESPN | 50.85 | 14.58 | 0.00584 |
| 10 | cg00212470 | 1 | 41301421 | KCNQ4 | 152.0 | 44.02 | 0.00621 |

Frequency 3 kHZ

|  | CpG | Chr | Pos | Gene | Estimate | SE | p |
| --- | --- | --- | --- | --- | --- | --- | --- |
| 1 | cg03660952 | 11 | 2644418 | KCNQ1 | 299.7 | 54.10 | 0.000247 |
| 2 | cg07124680 | 13 | 60587343 | DIAPH3 | -75.78 | 14.02 | 0.000299 |
| 3 | cg22102703 | 21 | 43346284 | C2CD2 | 92.79 | 17.69 | 0.000375 |
| 4 | cg18729973 | 21 | 43785785 | TFF1 | -89.37 | 18.00 | 0.000566 |
| 5 | cg07323754 | 21 | 39289266 | KCNJ6 | 48.10 | 10.43 | 0.000963 |
| 6 | cg08009711 | 21 | 43346184 | C2CD2 | 81.31 | 18.88 | 0.00154 |
| 7 | cg12161460 | 21 | 40818036 | LCA5L; SH3BGR | -219.9 | 53.93 | 0.00222 |
| 8 | cg19009644 | 3 | 10553211 | ATP2B2 | -82.21 | 20.54 | 0.00251 |
| 9 | cg27312626 | 21 | 39627877 | KCNJ15 | -65.65 | 17.15 | 0.00333 |
| 10 | cg09290941 | 14 | 31343685 | COCH | -66.80 | 17.64 | 0.00356 |

**Table 1d:** Top 10 CpG sites at audiometric frequency 3 kHZ , sorted by p- value.

Frequency 4 kHZ

|  | CpG | Chr | Pos | Gene | Estimate | SE | p |
| --- | --- | --- | --- | --- | --- | --- | --- |
| 1 | cg18729973 | 21 | 43785785 | TFF1 | -106.1 | 14.59 | 2.69E-05 |
| 2 | cg09290941 | 14 | 31343685 | COCH | -81.81 | 15.30 | 0.000324 |
| 3 | cg19009644 | 3 | 10553211 | ATP2B2 | -97.50 | 18.94 | 0.000432 |
| 4 | cg08009711 | 21 | 43346184 | C2CD2 | 92.79 | 18.77 | 0.000585 |
| 5 | cg19207856 | 21 | 38444952 | TTC3; PIGP | -113.6 | 26.26 | 0.00150 |
| 6 | cg11316868 | 21 | 38593754 | AP001432.14; DSCR9; TMPRSS3 | 85.67 | 20.49 | 0.00188 |
| 7 | cg02667467 | 17 | 72920176 | USH1G; OTOP2 | 110.6 | 27.58 | 0.00249 |
| 8 | cg22102703 | 21 | 43346284 | C2CD2 | 92.12 | 23.48 | 0.00285 |
| 9 | cg21063361 | 21 | 43255611 | PRDM15 | 41.97 | 10.95 | 0.00331 |
| 10 | cg15686157 | 11 | 2753885 | KCNQ1 | -104.7 | 27.92 | 0.00377 |

**Table 1e:** Top 10 CpG sites at audiometric frequency 4kHZ, sorted by p- value.

Frequency 8 kHZ

|  | CpG | Chr | Pos | Gene | Estimate | SE | p |
| --- | --- | --- | --- | --- | --- | --- | --- |
| 1 | cg22982528 | 21 | 40686185 | BRWD1 | -84.24 | 17.10 | 0.000598 |
| 2 | cg03660952 | 11 | 2644418 | KCNQ1 | 286.3 | 63.20 | 0.00109 |
| 3 | cg08895013 | 11 | 2468332 | KCNQ1 | -84.94 | 18.82 | 0.00112 |
| 4 | cg01176028 | 21 | 43653234 | ABCG1 | -49.29 | 12.83 | 0.00326 |
| 5 | cg07323754 | 21 | 39289266 | KCNJ6 | 45.07 | 12.04 | 0.00383 |
| 6 | cg15910264 | 11 | 2783880 | KCNQ1 | -74.90 | 20.72 | 0.00473 |
| 7 | cg02316713 | 21 | 43619559 | ABCG1 | 47.38 | 13.23 | 0.00500 |
| 8 | cg10939579 | 13 | 20768309 | GJB2 | -75.77 | 21.28 | 0.00518 |
| 9 | cg26776175 | 7 | 95064464 | PON2 | -114.8 | 32.74 | 0.00567 |
| 10 | cg25572105 | 7 | 95025955 | PON3 | 69.40 | 20.48 | 0.00690 |

**Table 1f:** Top 10 CpG sites at audiometric frequency 8kHZ, sorted by p- value.
